# CAF to the rescue! Potential and challenges of combination antifungal therapy for reducing morbidity and mortality in hospitalized patients with serious fungal infections

**DOI:** 10.1101/2024.07.10.24310241

**Authors:** Samantha E. Jacobs, Vishnu Chaturvedi

## Abstract

The global burden of invasive fungal disease (IFD) is substantial and escalating. Combination antifungal therapy (CAF) may improve patient outcomes by reducing development of resistance, improving drug penetration and rate of fungal clearance, and allowing for lower, less toxic antifungal drug doses; yet, increased cost, antagonism, drug-drug interactions, and toxicity are concerns. Clinical practice guidelines recommend antifungal monotherapy, rather than CAF, for most IFDs due to a lack of comparative randomized clinical trials. An examination of the existing body of CAF research should frame new hypotheses and determine priorities for future CAF clinical trials. We performed a systematic review of CAF clinical studies for invasive candidiasis, cryptococcosis, invasive aspergillosis, and mucormycosis. Additionally, we summarize findings from animal models of CAF and assess laboratory methods available to evaluate CAF efficacy. Future CAF trials should be prioritized according to animal models showing improved survival and observational clinical data supporting efficacy and safety.

## Introduction

### Continued dismal outcomes for patients with serious fungal infections

The global burden of invasive fungal disease (IFD) is substantial, causing approximately 1.5 million deaths per year, and escalating due to a growing population of immunosuppressed individuals, global health events such as the COVID-19 pandemic, and improvements in molecular diagnostics leading to increased reporting of previously unrecognized or undiagnosed IFD.(1–6)

IFDs have considerable economic impact in terms of direct medical costs, productivity loss, and pre-mature deaths.(7) In 2019, the estimated economic burden of fungal disease in the U.S. was $11.5 billion, and the mean cost of an inpatient visit associated with fungal disease diagnosis was more than twice that of the average inpatient stay. Within the healthcare system, IFDs lead to increased length of stay and mortality, and inappropriate empirical antimicrobial therapy.(8–10) Disparate mortality rates are observed across continents and countries. For example, 30-day all-cause mortality in patients with candidemia is approximately 30% in Europe and the U.S. and 60-72% in South Africa and Brazil.(11)

### Potential and challenges of combination antifungal therapy

Given the considerable individual and public health burden of IFDs, emphasis on research pertaining to antifungal therapeutics is essential. Except for cryptococcal meningitis, current clinical practice guidelines recommend antifungal monotherapy for most IFDs; yet, morbidity and mortality remains unacceptably high.(12–19) Combination antifungal therapy (CAF) is one putative strategy to reduce risk and improve outcomes. Potential reasons to pursue CAF for IFDs are: (1) To broaden the spectrum of activity for empirical therapy, (2) to enhance the rate of killing through antifungal synergy/ increased potency, (3) to delay or prevent development of resistance (4) to reduce antifungal toxicity by using lower drug dosages (5) or to enhance the effectiveness of older antifungals, and (6) to treat breakthrough infections.(20–23)

CAF may be particularly useful in difficult clinical situations, e.g., infections due to highly resistant pathogens or biofilm formation, or in tissues with compromised drug penetration. However, some potentially harmful effects of CAF include reduced fungal killing (antagonism), drug-drug interactions, increased toxicity, and increased cost.

The concept of combining drugs with different mechanisms of action to treat a disease is a longstanding practice supported by rigorous randomized controlled trials (RCTs) for other complex and severe infections including HIV and tuberculosis, as well as oncologic diseases.(24–27) Unfortunately, multiple challenges to conducting large-scale RCTs exist for patients with IFDs including cost, the significant time to conduct the study, particularly for rare fungi (e.g., Mucorales), and difficulty enrolling medically complex and/or critically ill patients.(28) Due to a lack of RCTs comparing CAF to monotherapy, CAF is reserved for salvage therapy or specific severe disease manifestations, e.g., *Candida* endocarditis, in U.S. and international guidelines.(13)

To frame new hypotheses and carefully inform clinical trial design, it is important to examine and build upon the existing body of preclinical research and human studies. We perform a systematic review of CAF clinical studies for the most prevalent and dangerous fungal pathogens, *Candida* spp., *Cryptococcus neoformans*, *Aspergillus* spp., and the Mucorales. We further summarize findings from experimental research and assess laboratory methods available to evaluate CAF efficacy. Table 1 provides a “bird’s eye view” qualitative summary of the evidence for CAF.

**Table 1.** Bird’s eye view: Summary of clinical and experimental evidence supporting combination antifungal therapy for the most prevalent and dangerous fungal pathogens.

| Disease | Qualitative Evidence Summary |
| --- | --- |
| Invasive Candidiasis | Single RCT of FLC vs. FLC + dAmB published in 2003 trended toward improved success in combination arm<br>Experimental models of systemic *C. albicans* and *C. glabrata* infection primarily support polyene-echinocandin combination<br>Paucity of experimental and clinical data investigating CAF for azole- and echinocandin-resistant *Candida* spp. and *C. auris*<br>No prospective trials incorporating echinocandins for CAF have been conducted |
| Cryptococcosis | Multiple large RCTs in HIV/AIDS-associated cryptococcal meningitis demonstrate that induction CAF with AmB + 5-FC leads to improved microbiologic and clinical outcomes compared to monotherapy<br>Single-dose L-AmB 10 mg/kg + 14 days x 5-FC and FLC now first-line regimen in resource-limited settings<br>No prospective trials in non-HIV/AIDS immunocompromised hosts, nor in non-CNS manifestations of cryptococcosis |
| Invasive Aspergillosis | Single large multination RCT of VOR + AFG vs. VOR alone trended toward improved 6-week mortality in HM patients with IA (P=0.087)<br>Azole-echinocandin combinations strongly supported by experimental models of IA<br>Experimental studies of azole-polyene combinations primarily demonstrate antagonism; very limited clinical data show no difference compared to AmB alone<br>Prospective clinical data to inform CAF for azole-resistant *Aspergillus* spp. are lacking |
| Mucormycosis | Mixed results of azole-polyene CAF from experimental data; polyene-echinocandin CAF *in vivo* synergy may be species-specific<br>No prospective trials of CAF exist; largest retrospective analysis of CAF including AmB, POS, ISA and echinocandins found similar 6-week survival compared to monotherapy |

5-FC, flucytosine; AFG, anidulafungin; CAF, combination antifungal therapy; CNS, central nervous system; dAmB, deoxycholate amphotericin B; FLC, fluconazole; ISA, isavuconazole; L-AmB, liposomal amphotericin B; POS, posaconazole; RCT, randomized clinical trial; VOR, voriconazole

### Clinical studies of combination antifungal therapy

#### Systematic review methods

A systematic review to identify clinical studies of CAF for invasive candidiasis (IC), cryptococcosis, invasive aspergillosis (IA), and invasive mucormycosis was conducted in accordance with the PRISMA guidelines.(29) PubMed was searched for English-language publications using the following terms “antifungal combination therapy” or “combination antifungal therapy” and “candida” or “candidiasis”, “cryptococcus” or “cryptococcosis” or “cryptococcal”, “aspergillus” or “aspergillosis”, and “mucorales” or “mucormycosis” or “zygomycetes” or “zygomycosis” (Appendix). We limited our search to manuscripts published between January 1, 2003 (coinciding with the year of U.S. FDA approval of voriconazole) and February 1, 2023. Reference lists from the included studies were additionally screened to identify potentially relevant evidence. We included studies whose primary aim was to evaluate the CAF efficacy for proven or probable IFD according to consensus definitions; fungal speciation was not required.(30, 31) Papers were excluded if they were literature reviews or meta-analyses, if they did not specify antifungal drug dosages, or if they evaluated combinations including non-systemic antifungal agents, e.g., topical or aerosolized formulations, or non-antifungal chemical compounds.

#### Systematic review results

The PRISMA flowcharts in the Appendix provide a detailed breakdown of the results of the primary search for each of the four IFDs. From 954 publications evaluated, 104 articles met the inclusion criteria. These included 21 CAF papers for IC, 17 for cryptococcosis, 45 for IA, and 21 for mucormycosis. Below, we focus our discussion on the findings from randomized trials and large observational studies for each pathogen. Details of each included manuscript, including study design, patient population, antifungal therapy, and patient-related outcomes, are provided in Supplementary Tables 1-4.

#### Invasive Candidiasis

Only one RCT of CAF for IC was identified in the systematic review, and the trial was conducted prior to the availability of echinocandins. The trial included 219 non-neutropenic patients who received fluconazole (FLC) 800 mg/d plus placebo vs. FLC plus deoxycholate amphotericin B (dAmB) 0.7 mg/kg/d for candidemia (*C. albicans* in 62%).(32) There was no difference between groups in the primary analysis of time-to-failure, nor in mortality, although overall success rates were higher in the combination vs. monotherapy arm (69% vs. 56%, respectively, *P*=0.043).

Further clinical data pertaining to CAF for IC was limited to 20 case reports and small case series, most often describing azole-echinocandin or polyene-echinocandin combinations used as salvage therapy. Four case reports and a series of 13 patients with peritoneal dialysis-associated *Candida* peritonitis described favorable outcomes in patients treated with combination dAmB or liposomal AmB (L-AmB) plus flucytosine (5-FC), a regimen still recommended for treatment of deep-seated *Candida* infections, e.g., meningitis and endocarditis, based on limited experimental and clinical data.(13, 33–37) This combination has shown synergy *in vitro* and is widely used for cryptococcal meningitis (CM).(38) Moreover, excellent 5-FC levels are achieved in cerebrospinal fluid (CSF).(39)

#### Cryptococcosis

The systematic review identified 12 RCTs and one prospective, non-randomized trial of induction therapy for CM, all in individuals with HIV/AIDS, as well as 3 retrospective studies and a case report in persons with other immunocompromising conditions. There are no prospective trials in persons without HIV, nor has CAF been evaluated for other manifestations of cryptococcosis, e.g., pulmonary disease.

Several trials established that the combinations of AmB plus 5-FC and AmB plus FLC both achieve improved clinical outcomes as compared to AmB alone.(40–42) However, fungicidal activity, as measured by the rate of reduction in CSF yeast CFU from serial quantitative cultures, is greater with AmB plus 5-FC as compared to AmB plus FLC.(40, 41)

Combination 5-FC and FLC is appealing due to its oral availability and low rates of nephrotoxicity, although prolonged 5-FC is associated with leukopenia.(43) In a pivotal trial in Africa (ACTA) including 721 patients, the all-oral induction regimen of FLC 1200 mg/d plus 5-FC 100 mg/kg/d was non-inferior to dAmB plus FLC or 5-FC for 7 or 14 days for the primary endpoint of 2-week mortality.(44) However, faster CSF clearance was observed with AmB plus 5-FC compared to FLC plus 5-FC.

L-AmB was investigated in a landmark trial, Ambition, in 844 individuals with CM in sub-Saharan Africa. A single 10 mg/kg dose of L-AmB plus 14 days of high-dose FLC and 5-FC was non-inferior to the WHO-recommended 1-week regimen of dAmB plus 5-FC and was associated with fewer adverse events.(45) Following the results of the ACTA and Ambition trials, as well as advocacy efforts, generic 5-FC is more widely available at a reduced cost and utilized in routine care.(46)

Three-drug combinations of AmB, 5-FC, and FLC have also been studied. While the triple therapy had the most potent fungicidal effect in a murine model of CM, it was not more efficacious than AmB plus 5-FC in humans.(40, 47)

Other triazole antifungal agents have excellent *in vitro* activity against *Cryptococcus* spp., and the combination of AmB and voriconazole (VOR) had similar early fungicidal activity as compared to AmB plus 5-FC and AmB plus FLC in a small randomized trial.(48, 49) However, VOR, isavuconazole (ISA), and posaconazole (POS) have more side effects and drug-drug interactions, and higher costs than FLC, thus limiting their utility for a disease that is most prevalent in resource-limited settings.

#### Invasive Aspergillosis

Two randomized trials, 10 observational studies, and 33 case reports/series met inclusion criteria in the systematic review of CAF for IA.

Prior to widespread use of VOR as standard of care, prospective studies primarily investigated polyene-echinocandin combination. In the randomized, open-label Combistrat trial including 30 patients with hematologic malignancy (HM) and IA, those receiving L-AmB 3 mg/kg/d plus caspofungin (CAS) had higher favorable response rate (67%) and less acute kidney injury than those receiving high-dose L-AmB 10 mg/kg/d alone (27%, P=0.028 for comparison) for primary therapy.(50) However, L-AmB ≥ 7.5 mg/kg/d plus CAS combination led to poorer clinical outcomes compared to POS suspension for salvage therapy in a single center compassionate use trial.(51) Further clinical trials with polyene-based combination therapy have not been pursued in the past two decades given a lack of compelling preclinical data, high rates of AmB-associated nephrotoxicity, and studies confirming improved IA-related mortality with VOR.(52, 53)

Observational studies of echinocandin-triazole combination therapy for IA have had inconsistent results depending on the indication and study population. Marr and colleagues compared VOR plus CAS to VOR alone in 47 patients with HM and hematopoietic cell transplant (HCT) recipients with IA who had failed initial therapy with AmB. Improved 3-month survival was observed in the combination group compared to monotherapy (HR, 0.42; 95% CI, 0.17–1.1; *P*=0.048), independent of other variables associated with prognosis.(54) However, in a retrospective study spanning 12 years (1998–2010), VOR plus CAS did not achieve better outcomes than VOR alone, as primary or salvage therapy.(55) In solid organ transplant recipients, compared to a historical cohort receiving L-AmB alone, VOR plus CAS for primary therapy of IA led to similar overall survival at Day 90 (67.5% combination vs. 51% L-AmB monotherapy controls; *P*=0.117). (56)

To date, one RCT has evaluated echinocandin-triazole combination for IA. This multicenter, multinational study randomized patients with HM or HCT recipients to VOR plus anidulafungin (AFG) versus VOR alone for primary therapy of proven or probable IA.(57) Amongst 277 patients, there was no difference in 6-week mortality (combination: 19.4%, monotherapy: 27.5%; *P*=0.087). However, in a post-hoc subgroup analysis of probable IA cases (n=218), 6-week mortality was lower in the combination arm (15.7% vs. 27.3% monotherapy; *P*=0.037). A limitation of this trial is that statistical power was lower than expected due to higher than expected mortality in both treatment arms.

Polyene-triazole combination therapy has received relatively little study in humans given the preclinical data demonstrating variable synergistic interactions.(58) The only comparative data are from a retrospective study in which similar 12-week survival was observed among 49 patients receiving L-AmB 3 mg/kg/d plus VOR or POS vs. L-AmB plus CAS vs. VOR plus echinocandin.(59)

Taken together, these single studies have failed to decisively establish a benefit to CAF for IA; however, limitations in study design may preclude definitive determination.

#### Mucormycosis

Twenty-one manuscripts, including 6 retrospective studies and 15 case reports/series were included in the systematic review. A 2008 retrospective study by Reed and colleagues was one of the first to suggest a benefit to polyene-echinocandin combination therapy for primary treatment of rhino-orbital-cerebral mucormycosis (ROCM). Amongst 41 patients with ROCM (83% diabetic) between 1994-2006 at two centers, 100% (7 of 7) of patients receiving lipid-complex AmB (ABLC) or L-AmB plus CAS were alive and not in hospital care 30 days after hospital discharge vs. 45% (15/34) of patients on dAmB or a lipid formulation alone (*P*=0.02).(60) In a multivariate analysis, only receipt of combination therapy was significantly associated with improved outcome (OR, 10.9; *P*=0.02).

Kyvernitakis and colleagues performed the largest analysis to date in a retrospective study including 106 HM and HCT patients with mucormycosis at a single U.S. cancer center between 1994-2014.(61) For initial therapy, 44% of patients received monotherapy (L-AmB 87%, POS 13%) and 56% of patients received CAF (L-AmB + echinocandin, 46%; L-AmB + POS, 27%; triple therapy, 27%), highlighting the frequency with which CAF is employed in clinical practice. Using a propensity-score adjusted analysis, they found a similar 6-week survival between patients receiving initial monotherapy vs. combination therapy (56% vs. 60%; *P*=0.71).

An important feature of the abovementioned studies is the relatively long study period spanning over 10 years, which may introduce biases pertaining to changes in fungal diagnostics, supportive care practices, and availability of specific antifungal agents and formulations. An additional limitation to observational studies of IFD treatment is the use of different dosages of antifungal agents, which may affect efficacy and toxicity.

### Animal studies of combination antifungal therapy

In the absence of large-scale RCTs, animal models of IFDs have several strengths. They allow for pharmacokinetics of the administered drugs, tissue burden, and rate of clearance to be assessed. Researchers can integrate host factors and study a range of doses and dose combinations. Finally, resistant species may be studied; whereas, enrolling patients with rare and highly resistant fungal pathogen into clinical trials is quite challenging. With the implementation of ARRIVE (Animal Research: Reporting In Vivo Experiments) and ARRIVE 2.0 guidelines on the design, conduct, and reporting of animal studies, it is reasonable to conclude that animal studies with IFD and CAF will be able to bridge the quality gap with RCTs.(62–64)

#### Candida spp

Studies of CAF in experimental models of infections due to *Candida* spp. have yielded mixed results depending on the species, drugs, and methodology. For example, the combination of FLC plus CAS for murine candidemia (*C. albicans*) yielded no change in tissue fungal burden as compared to FLC monotherapy; whereas POS plus CAS led to improved survival compared to either drug alone.(65, 66)

An *in vivo* study by Louie and colleagues further illustrates the complexity of antifungal interactions.(67, 68) In a rabbit model of *Candida albicans* endocarditis and pyelonephritis, both AmB monotherapy and sequential treatment with AmB alone followed by AmB plus FLC rapidly sterilized kidneys and cardiac vegetations. In contrast, simultaneous AmB plus FLC, FLC monotherapy, and FLC followed by AmB were all slower to clear fungi from infected tissues.(68)

Polyene-echinocandin combination is the most frequently studied regimen in the experimental models of IC, particularly due to *C. glabrata*. Compared to monotherapy, AmB plus CAS or L-AmB plus CAS or micafungin (MFG) led to reduced tissue fungal burden in a murine model of systemic infection due to azole-resistant *C. albicans* and *C. glabrata* and in immunosuppressed mice with *C. glabrata* infection.(69–71)

#### Cryptococcus spp

*In vivo* studies of CAF for cryptococcosis laid the groundwork for subsequent RCTs in humans. In particular, AmB, FLC, and 5-FC, in different dosages and combinations, have been extensively studied in animal models of disseminated disease and meningitis. Schwarz and colleagues demonstrated that AmB plus 5-FC, as compared to monotherapy, led to improved survival and reduced brain tissue fungal burden in murine disseminated cryptococcosis due to 5-FC-susceptible or -resistant *C. neoformans*.(72) Other studies have found that FLC and AmB, whether given sequentially or combined, lead to greater antifungal activity than AmB alone.(73, 74) The role of high-dose FLC, with or without 5-FC, was further established in murine cryptococcal meningitis.(75)

Other triazoles have received lesser attention, either as monotherapy or combination therapy, in experimental models of cryptococcosis.(76)

#### Aspergillus spp

Preclinical studies of azole-echinocandin combinations for IA have garnered significant interest given the favorable safety profile of the latter group. The combinations of POS plus AFG and ISA plus MFG led to improved outcomes in neutropenic models of IA.(77, 78) This synergistic interaction between the echinocandin and the triazole is likely due to simultaneous inhibition of biosynthesis of 1,3-β-D-glucan in the fungal cell wall and ergosterol in the cell membrane. However, poorer outcomes observed with VOR plus high-dose AFG suggest that azole-echinocandin interactions may be concentration-dependent.(79)

Experimental models of IA treated with azole-polyene combinations have primarily demonstrated antagonism except for CNS disease.(79–82) Again, consideration of the drug mechanism of action may explain these observations. Antifungal azoles deplete the fungal cell membrane of ergosterol and thereby diminish the principal biochemical target of AmB.

Polyene-echinocandins combination did not show any benefit for experimental CNS aspergillosis or for invasive pulmonary aspergillosis in immunosuppressed murine models due to chronic granulomatous disease or glucocorticoids.(82–84)

#### Mucorales

Azole-polyene combinations for treatment of mucormycosis are frequently used in clinical practice, although preclinical data do not clearly establish a benefit. In neutropenic mice with pulmonary mucormycosis due to *R. delemar* or *M. circinelloides*, L-AmB plus ISA improved survival and reduced tissue fungal burden (lung and brain) compared to either drug alone.(85) However, in murine disseminated mucormycosis due to *R. oryzae*, L-AmB plus POS did not improve outcomes as compared to L-AmB monotherapy.(86, 87)

Echinocandins do not have *in vitro* activity against Mucorales in standard susceptibility tests, however, some agents of mucormycosis, including *Rhizopus oryzae*, express (1→3)-β-D-glucan synthase, which is the target enzyme for echinocandins.(88) Experimental studies of murine mucormycosis due to *R. oryzae* have found improved outcomes when combining lipid formulations of Amb plus echinocandins as compared to either agent alone.(89, 90) The benefit noted with the addition of echinocandins may be in part due to the class’ immunomodulatory effect on the activity of phagocytic cells.(91) However, no evidence of synergy nor antagonism was noted for the combination of ISA and MFG in neutropenic mice with pulmonary disease due to *R. delemar*.(92)

### Laboratory methods

There are no standard CLSI or EUCAST methods for *in vitro* testing of antifungal agents in combinations (93). Our literature analysis revealed numerous publications on laboratory testing of antifungal drugs in combinations against yeasts and molds (Supplementary Fig 1). Only a small selection of published studies included details consistent with the inoculum, inclusion of quality control strains, end-point reading, and interpretations of test results per CLSI or EUCAST methods for testing of molds and yeasts (94–112). Even publications describing combination testing with standard methods originated from single institutions and the reproducibility of the described method at other institutions remains unknown. The development of a consensus method with standardized parameters is imperative for the meaningful testing of antifungal combinations in the clinical laboratory. Towards this end, one of us conducted a series of multi-laboratory studies to define the parameters that would allow reproducible antifungal combination testing. We identified a *Candida krusei* QC strain, inoculum size, and 100% inhibition end-point as highly reproducible features for combination testing of *Candida* species and *Aspergillus fumigatus* among six laboratories (113–115). The result interpretations were consistent among laboratories using summation fractional inhibitory concentration indices (ΣFICI) (113, 116). Recent re-analysis of the data revealed significant pharmacodynamic interactions with Loewe additivity-based FICi range of 1–2 FIC and essential agreement with Bliss independence-based response surface analysis (116). Thus, laboratories now have access to a standardized method for *in vitro* testing of antifungal agents in combinations. Further progress is imminent when multi-laboratory studies are conducted with newer antifungal combinations and with other fungal pathogens. Similarly, there is an opportunity for device manufacturers to offer commercial combination panels for wider access to routine laboratory testing.

### Future prospects

Table 2 summarizes ongoing clinical trials of CAF, including those incorporating investigational and non-systemic (e.g., aerosolized) antifungal agents. Promising drugs in clinical phases of development with novel mechanisms of action include fosmanogepix, olorofim, and ibrexafungerp.(117) These agents offer hope to expand our antifungal armamentarium, particularly against resistant yeasts and molds, yet will face the same environmental and host pressures for emergence of resistance over time. As such, drug development research should investigate these compounds in combinations and even look towards an antifungal “polypill” that might improve patient adherence and lessen healthcare costs.(23, 118)

**Table 2:**
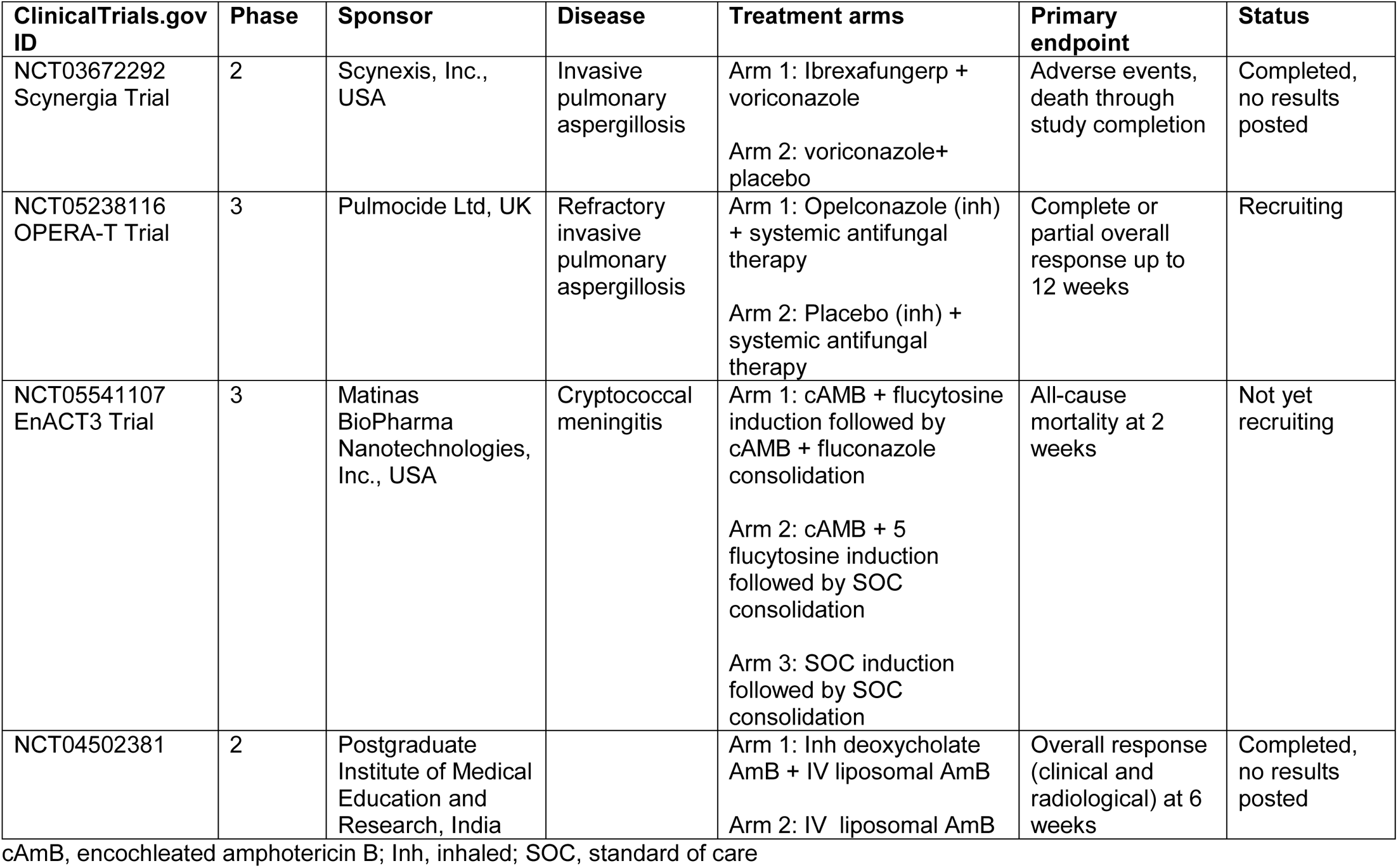
Ongoing clinical trials of combination antifungal therapy for invasive fungal diseases.

While our review focused on a combinatorial strategy involving antifungal drugs, a particularly novel and emerging area of research is the role of cancer immunotherapies as adjunctive therapy for IFDs. Evidence that fungi induce activation of checkpoint pathways has led to preclinical data demonstrating that blockade of the Programmed Cell Death Protein 1 (PD-1) and Cytotoxic T-Lymphocyte Antigen 4 pathways augments antifungal immunity and improves outcomes.(119) Thus far, clinical data are limited to case reports for invasive mold diseases.(120, 121) Besides providing support towards future interventional trials, such investigations of immunomodulatory therapies to combat IFDs highlight the need to understand the role of host immune responses in the outcomes of IFDs when designing and interpreting experimental and clinical studies of CAF.

## Conclusion

Our systematic review of the literature highlights the paucity of rigorous clinical investigations of CAF for invasive candidiasis and mucormycosis; whereas, multiple RCTs establish the superiority of polyene-5-FC CAF for cryptococcosis and one large RCT of azole-echinocandin CAF for IA suggests a benefit in patients with hematologic malignancy and probable IA. Given the challenges to conducting large prospective RCTs for most IFDs, it is reasonable to prioritize future trials of CAF based upon animal models demonstrating improved survival and observational clinical data supporting efficacy and safety. Recent multi-laboratory investigations demonstrated the feasibility of a standardized method of *in vitro* testing of antifungal combinations for *Candida* species that should be further investigated with other pathogen and drug combinations.

## Data Availability

All data produced in the present study are available upon reasonable request to the authors.

## Author contributions

S.J. and V.C: Conceptualization, systematic literature review, writing of original draft, and review and editing.

## Conflict of interest

The authors have no conflicts of interest to declare.

## Funding

None.

## Patient consent statement

Not applicable.

## Notes

### Competing Interest Statement

The authors have declared no competing interest.

### Funding Statement

This study did not receive any funding.

