## Supplemental Tables S1-4 for "CAF to the rescue! Potential and challenges of combination antifungal therapy for reducing morbidity and mortality in hospitalized patients with serious fungal infections"

**Supplementary data**

**Table S1. Systematic review of publications evaluating combination antifungal therapy for invasive candidiasis, 2003-2023**

| 1st author, year | Study design | Study period | Study population | N | Antifungal therapy† | Treatment indication | Results - Primary endpoint | Safety outcomes | Pediatric patients | Comments |
|---|---|---|---|---|---|---|---|---|---|---|
| Mikamo, 2003[1] | Case report | 2002 | 29F solid tumor | 1 | FLC 400 mg/d + dAmB 0.5 mg/kg/d | Primary therapy, *C. glabrata* peritonitis | Resolved | None | N | |
| Rex, 2003[2] | Prospective, randomized, blinded, multicenter | 1995-1999 | Non-neutropenic hospitalized | 219 | FLC 800 mg/kg + PBO vs FLC 800 mg/d + dAmB 0.7 mg/kg/d | Initial therapy for candidemia | No difference in time-to-failure between groups, P = 0.08 | Renal dysfunction in 3% (FLC + PBO) vs. 23% (FLC + dAmB), P < 0.001 | Yes, >= 13 years-old | PBO or AmB component only for first 5-8 days of therapy; overall success rates 56% of FLC + PBO vs. 69% of FLC + dAMB (P = 0.043) while the bloodstream infection failed to clear in 17% and 6% of subjects, respectively (P = 0.02) |
| Pelletier, 2005[3] | Case report | NR | 49F intestinal perforation | 1 | ABLC 5 mg/kg/d + 5-FC 2 g BID, followed by VOR + 5-FC | Salvage therapy, disseminated *C. krusei* including CNS | Resolved | NR | N | Progressive infection on CAS monotherapy for initial treatment |
| Sim, 2005[4] | Case report | 2002 | 53M ALL | 1 | FLC 600 mg/d + CAS | Salvage therapy, *C. tropicalis* septic arthritis | Resolved | none | N | |
| Lopez-Ciudad, 2006[5] | Case report | NR | 59F cirrhosis | 1 | VOR + CAS | Salvage therapy, *C. parapsilosis* mural endocarditis | Resolved | None | N | VOR added to CAS after 72 hours of initial therapy; surgical mangement deferred due to comorbidites |

| | | | | | | | | | | |
|---|---|---|---|---|---|---|---|---|---|---|
| Olver, 2006[6] | Case report | NR | 24M ALL | 1 | L-AmB 5 mg/kg/d + CAS | Salvage therapy, *C. krusei* bloodstream infection | Resolved | none | N | |
| Wong, 2008[7] | Retrospective case series | 2000-2005 | Continuous ambulatory peritoneal dialysis | 13 | dAmB 0.75-1 mg/kg/d + 5-FC 1 g/d | Primary therapy, *Candida* peritonitis | Microbiologic clearance in 10 (76.9%), 2 (15.4%) deaths related to peritonitis | Due to elevated LFTs, 5-FC stopped early in 2 patients, and both 5-FC and dAmB stopped early in 1 patient | N | Peritoneal catheter removed in 9 patients; outcomes also compared to historical control group of 14 patients (12 treated with antifungal monotherapy) that had higher peritonitis-related mortality (4/14, 28.5%) |
| Filippi, 2009[8] | Case report | NR | Neonate | 1 | FLC 6 mg/kg q 48 h + L-AmB 3 mg/kg/d + CAS 1 mg/kg/d | Salvage therapy, *C. albicans* bloodstream infection | Resolved | None | Y | |
| Bland, 2009[9] | Case report | 2008 | 55F rheumatoid arthritis | 1 | MFG 100 mg/d + FLC 400 mg/d x 8 weeks, then FLC alone | Primary therapy, *C. albicans* prosthetic joint infection | Initial clinical and microbiological resolution, then clinical relapse on FLC monotherapy requiring hardware removal and reinitiation of combination therapy | None | N | |
| Turan, 2011[10] | Retrospective case series | 2006-2009 | Neonate | 6 | VOR 6 mg/kg q 8 h + L-AmB 5 mg/kg/d | Salvage therapy, *Candida* sepsis | Resolved | Elevated LFTs (n=4) | Y | Primary therapy included FLC alone, FLC + dAmB, or FLC + L-AmB |

| | | | | | | | | | | |
|---|---|---|---|---|---|---|---|---|---|---|
| Al-Sweih, 2015[11] | Case report | NR | Neonate | 1 | L-AmB 8.5 mg/kg/d + CAS 2.5 mg/kg/d | Salvage therapy, *C. fermentati* bloodstream infection | Resolved | None | Y | |
| Reyes, 2015[12] | Case report | NR | 36F ovarian cancer | 1 | AFG 100 mg/d + VOR 200 mg BID | Initial therapy, C. parapsilosis endocarditis | Resolved | None | N | Fluconazole suppression following 60 days x AFG + VOR |
| Mpakosi, 2016[13] | Case report | NR | Neonate | 1 | L-AmB 7 mg/kg/d + MFG 10 mg/kg/d | Salvage therapy, *C. pulcherrima* bloodstream infection | Resolved | none | Y | Checkerboard broth microdilution, synergy between AmB and MFG (FIC 0.375) |
| Al-Sweih, 2017[14] | Case report | NR | Neonate | 1 | L-AmB 8.5 mg/kg/d + CAS 2.5 mg/kg/d | Salvage therapy, *C. conglobata* bloodstream infection | Resolved | none | Y | |
| Tsakiri, 2018[15] | Case report | NR | Neonate | 1 | L-AmB 5 mg/kg/d + MFG 10 mg/kg/d + VOR 9 mg/kg q 12 h + intrathecal AmB 0.025 mg x 1 dose | Salvage therapy, *C. glabrata* meningitis with ventricular access device | Initial CSF sterilization, then relapse 34 days later that resolved after removal of hardware | none | Y | |
| Ahuja, 2019[16] | Case series | NR | Prosthetic valve endocarditis | 2 | Case 1: MFG 150 mg/d + FLC 400 mg/d x 12 weeks, then FLC 200 mg/d suppression; Case 2: L-AmB 5 mg/kg/d + 5-FC 100 mg/kg/d + FLC 400 mg/d, then MFG 150 mg/d + 5-FC + FLC, then 5-FC + FLC suppression | Salvage therapy, *C. parapsilosis* PVE | Resolved | AKI on L-AmB | N | |

| | | | | | | | | | | |
|---|---|---|---|---|---|---|---|---|---|---|
| Keuning, 2019[17] | Case report | 2018 | 72F rheumatoid arthritis | 1 | VOR + MFG 200 mg/d x 4 weeks, then VOR 300 mg BID alone | Primary therapy, *C. parapsilosis* prosthetic joint infection | Resolved | Elevated LFTs attributed to MFG | N | Two-stage exchange arthroplasty with antifungal spacer (AmB) performed |
| Wee, 2021[18] | Case report | NR | Elderly F hemorrhagic stroke | 1 | L-AmB 5 mg/kg/d + 5-FC 100 mg/kg/d x 14 days, then FLC 12 mg/kg/d x 35 days | Primary therapy, *C. glabrata* meningitis | Resolved | None | N | |
| Jacobs and Jacobs, 2022[19] | Case report | 2020 | 28F multivisceral transplant | 1 | VOR + MFG 150 mg/kg/d + ibrexafungerp 750 mg daily | Salvage therapy, *C. auris* peritonitis | Died | Leukopenia related to 5-FC | N | |
| Odysseos, 2022[20] | Case report | NR | 52F liver transplant | 1 | L-AmB 6 mg/kg/d + ISA | Salvage therapy, *C. glabrata* bloodstream infection and bilomas | Resolved | None | N | |
| Sharma, 2023[21] | Case report | NR | 55M rheumatic heart disease | 1 | CAS 150 mg/d + FLC 400 mg/d x 28 days, then FLC 400 mg/d | Primary therapy, *C. parapsilosis* prosthetic valve endocarditis | Died | None | N | Valve surgery deferred due to cachexia |

5-FC, flucytosine; AFG, anidulafungin; AKI, acute kidney injury; ALL, acute lymphoblastic leukemia; CAS, caspofungin; CSF, cerebrospinal fluid; dAmB, deoxycholate amphotericin B; FLC, fluconazole; L-AmB, liposomal amphotericin B; LFTs, liver function tests; MFG, micafungin; NR, not reported; PBO, placebo; PVE, prosthetic valve endocarditis; SAE, serious adverse event

† Antifungal dosages unless otherwise specified: CAS 70 mg x 1 dose followed by 50 mg/d, VOR 6 mg/kg q 12 h x 2 doses followed by 4 mg/kg q 12 h, ISA 372 mg q 8 h x 6 doses followed by 372 mg/d

**Table S2. Systematic review of publications evaluating combination antifungal therapy for cryptococcosis, 2003-2023**

| 1st author, year | Study design | Study period | Study population | N | Antifungal therapy† | Treatment indication | Results - Primary endpoint | Safety outcomes | Pediatric patients | Comments |
|---|---|---|---|---|---|---|---|---|---|---|
| Brouwer, 2004[22] | Randomized trial | 2002, Thailand | HIV/AIDS | 64 | dAmB 0.7 mg/kg/d alone vs. dAmB + 5-FC 100 mg/kg/d vs. dAmB + FLC 400 mg/d vs. triple therapy x 14 days, followed by FLC 400 mg/d | Induction therapy, CM | Higher EFA with dAmB + 5-FC vs. dAmB alone (P = 0.0006), dAmB + FLC (P = 0.02), or triple therapy (P = 0.02) | No study drug withdrawal due to side effects during first 2 weeks | N | No mortality difference across groups |
| Bicanic, 2008[23] | Randomized trial | 2005-2006, South Africa | HIV/AIDS | 64 | dAmB 0.7 mg/kg/d + 5-FC 100 mg/kg/d vs. dAmB 1 mg/kg/d + 5-FC x 14 days, followed by FLC 400 mg/d | Induction therapy, CM | Higher EFA with dAmB 1 mg/kg/d + 5-FC than dAmB 0.7 mg/kg/d + 5-FC (P = 0.05) | No difference in renal impairment between groups | N | No mortality difference between groups |
| Milefchik, 2008[24] | Randomized trial | 1991-1994 | HIV/AIDS | 89 | FLC 800mg or 1200 mg or 1600 mg or 2000 mg/d x 10 weeks +/- 5-FC 100 mg/kg/d x 4 weeks | Induction therapy, CM | Alive with negative CSF culture by week 10: FLC 800 mg alone (11%) or plus 5-FC (75%), FLC 1200 mg alone (37%) or plus 5-FC (87%), FLC 1600 alone (62%) or plus 5-FC (62%), FLC 2000 mg alone (62%) or plus 5-FC (75%) | Elevated LFTs and granulocytopenia, respectively in 2% and 7% FLC alone and 7% and 18% combination groups | N | |

| | | | | | | | | | | |
|---|---|---|---|---|---|---|---|---|---|---|
| Pappas, 2009[25] | Randomized trial, phase 2 | 2005-2007, U.S. and Thailand | HIV/AIDS | 143 | dAmB 0.7 mg/kg/d alone vs. dAmB 0.7 mg/kg/d + FLC 400 mg/d vs. dAmB 0.7 mg/kg/d + 800 mg/d x 14 days, followed by FLC alone at randomized dosage | Induction therapy, CM | No difference in rates of severe toxicity between groups; higher rate of successful outcome in dAmB + FLC 800 mg group (53.7%) vs. dAmB alone (41.3%) and dAmB + FLC 400 mg (27.1%) | Most common toxicities hypokalemia and hypomagnesemia | N | Primary efficacy endpoint = composite of CSF sterilization, stable neurological function, and survival at day 14 |
| Nussbaum, 2010[26] | Randomized trial | 2008, Malawi | HIV/AIDS | 41 | FLC 1200 mg/d alone vs. FLC + 5-FC 100 mg/kg/d x 14 days, followed by FLC 800 mg/d | Induction therapy, CM | Higher EFA in combination arm compared to monotherapy (P = <0.001) | More grade 3 or 4 neutropenia in combination arm | N | Fewer deaths in combination arm at 2 weeks (10% vs. 37%) and at 10 weeks (43% vs. 58%) |
| Jackson, 2012[27] | Randomized trial | 2009-2010, Malawi | HIV/AIDS | 40 | FLC 1200 mg/d + dAmB 1 mg/kg/d vs. FLC + 5-FC 100 mg/kg/d + dAmB 1 mg/kg/d x 14 days, followed by FLC 800 mg/d | Induction therapy, CM | Higher EFA in triple combination arm compared to FLC + dAmB (P = 0.03) | | N | No difference in mortality between groups |
| Loyse, 2012[28] | Randomized trial | 2006-2008, South Africa | HIV/AIDS | 80 | dAmB 0.7-1 mg/kg/d + 5-FC 100 mg/kg/d vs. dAmB + FLC 800 mg/kd vs. dAmB + FLC 600 mg/d vs. dAmB + VOR 300 mg BID x 14 days, followed by FLC 400 mg/d | Induction therapy, CM | No difference in EFA among groups | No significant difference in grade 3 and 4 AEs among groups | N | |

| | | | | | | | | | | |
|---|---|---|---|---|---|---|---|---|---|---|
| Muzoora, 2012[29] | Prospective, open-label, compared to historical cohort | 2008-2009, Uganda | HIV/AIDS | 90 | dAmb 1 mg/kg/d x 5 days + FLC 1200 mg/d x 14 days (n=30) vs. historical cohort treated with FLC 800 mg/d or 1200 mg/d alone x 14 days (n=60) | Induction therapy, CM | More rapid EFA in combination group than FLC 800 mg or 1200 mg groups ($P < 0.001$ for both) | No grade 4-5 anemia or elevated Cr in combination group | N | Overall mortality 28% at 10 weeks in combination group |
| Day, 2013[30] | Randomized trial | 2004-2010, Vietnam | HIV/AIDS | 299 | dAmB 1 mg/kg/d x 4 weeks vs. dAmB + 5-FC 100 mg/kg/d x 2 weeks vs. dAmB + FLC 400 mg BID x 2 weeks | Induction therapy, CM | Fewer deaths in dAmB + 5-FC arm compared to dAmB alone arm at day 14 ($P = 0.08$) and day 70 ($P = 0.04$); no difference in mortality between dAmB alone and dAmB + FLC arms; higher EFA in dAmB + 5-FC arm compared to both dAmB alone and dAmB + FLC arms | common in combination therapy groups compared to dAmB alone | N | First study to show a mortality benefit with combination therapy |
| Vaidhya, 2015[31] | Randomized trial | 2012-2014, India | HIV/AIDS | 84 | dAmB 0.7-1 mg/kg/d alone vs. dAmB + FLC 400 mg/d x 14 days | Induction therapy, CM | No difference in rates of fever, headache, neck stiffness between groups at 14 days, but greater improvement in altered sensorium and focal neurologic deficits and lower rates of CSF culture positivity at day | NR | N | |

| | | | | | | | | | | |
|---|---|---|---|---|---|---|---|---|---|---|
| | | | | | | | 14 in combination arm vs. monotherapy. | | | |
| Molloy, 2018[32] | Randomized trial (ACTA) | 2013-2016, Africa | HIV/AIDS | 678 | FLC 1200 mg/d + 5-FC 100 mg/kg/d x 2 weeks vs. dAmB 1 mg/kg/d + FLC 1200 mg/kg OR 5-FC 100 mg/kg/d x 1 week followed by FLC 1200 mg/d vs. dAmB 1 mg/kg/d + FLC OR 5-FC x 2 weeks | Induction therapy, CM | Similar mortality in the oral-regimen, 1-week dAmB, and 2-week dAmB groups: 18.2%, 21.9%, and 21.4%, respectively, at 2 weeks, and 35.1%, 36.2%, and 39.7%, respectively, at 10 weeks | Grade 4 anemia and grade 3-4 serum creatinine increase more common in dAmB groups compared to oral-regimen group | N | Lowest 10-week mortality in 1-week dAmB + 5-FC group (24.1%;95% CI, 16.2-32.1); more rapid CSF sterilization in dAmB + 5-FC groups compared to dAmB + FLC and to oral-regimen groups |
| Jandhyala, 2019[33] | Case report | NR | Critical illness | 1 | L-AmB 5 mg/kg/d + 5-FC 50 mg/kg/d x 2 weeks, followed by FLC 800 mg/d | Primary therapy, disseminated cryptococcosis (blood and lungs) | Survived | NR | N | |

| | | | | | | | | | | |
|---|---|---|---|---|---|---|---|---|---|---|
| Jarvis, 2019[34] | Randomized trial | 2014-2016, Botswana and Tanzania | HIV/AIDS | 80 | L-AmB 10 mg/g day 1 only vs. 10 mg/kg day 1 and 5 mg/kg day 3 vs. L-AmB 10 mg/kg day 1 and 5 mg/kg days 3 and 7 vs. L-AmB 3 mg/kg/d x 14 days (control arm). All groups received FLC 1200 mg/d x 14 days. | Induction therapy, CM | Non-inferior EFA in all short-course L-AmB arms compared to control arm. | Low rates of grade ≥ 3 clinical and laboratory AEs overall and no difference between treatment arms | N | No significant difference in 2-week and 10-week mortality between treatment arms. |
| Li, 2019[35] | Retrospective cohort study | 2010-2017, China | Non-HIV, non-transplant | 90 | dAmB 0.57-0.98 mg/kg/d + 5-FC 55.75 +/- 14.30 mg/kg/d (n=43) vs. FLC 400-800 mg/d + 5-FC 57.64 +/- 13.34 mg/kg/d (n=47) | Induction therapy, CM | No difference in rate of CSF clearance at 12 weeks in dAmB (74.4%) vs. FLC (70.2%) groups, P = 0.814 | Fewer AEs in FLC vs dAmB group. | N | At baseline, higher CSF cryptococci burden in FLC group. No difference in EFA, treatment response, or mortality (overall 3.3%) between groups. |
| Jarvis, 2022[36] | Randomized trial | 2018-2012, Africa | HIV/AIDS | 814 | L-AmB 10 mg/kg x 1 dose + 5-FC 100 mg/kg/d and FLC 1200 mg/d vs. dAmB 1 mg/kg/d + 5-FC 100 mg/kg/d x days followed by FLC 1200 mg/d x 7 days (control arm) | Induction therapy, CM | 10-week mortality non-inferior in L-AmB arm (24.8%) vs. control arm (28.7%). | Fewer life-threatening AEs in L-AmB arm (21.7%) vs. control arm (30.1%; P = 0.005) and less grade 3-4 anemia in L-AmB arm than control arm (P < 0.001) | N | No difference in EFA between treatment arms |

| | | | | | | | | | | |
|---|---|---|---|---|---|---|---|---|---|---|
| Takazono, 2022[37] | Retrospective cohort study | 2010-2017, Japan | Hospitalized patients without HIV (malignancy [29%], steroid usage [63.1%], collagen diseases [22.9%]) | 363 | L-AmB alone (median dose 3.49 mg/kg/d, IQR 2.51-4.64), n=146 patients vs. L-AmB (median dose 3.75 mg/kg/d, IQR 2.78-4.71) + 5-FC, n=217 patients | Induction therapy, CM | No difference in mortality 14 days after diagnosis in L-AmB alone vs combination group (11% vs. 6%, respectively; HR 0.5775, 95% CI 0.28-1.21, P = 0.1) | NR | N | 5-FC dose not specified |
| Zhao, 2022[38] | Retrospective cohort study | 2013-2018, China | Persons without HIV (steroid usage [31.6%], diabetes mellitus [23.7%], liver cirrhosis [7.9%]) | 38 | FLC 800 mg/d + 5-FC 78-100 mg/kg/d (n=27) vs. FLC 800 mg/d alone (n=11) | Initial therapy, CM | Successful response to initial therapy in 77.8% (combination arm) and 72.7% (monotherapy arm)† | No difference in rates of adverse drug reactions between groups; rate of leukopenia not reported | N | Median duration of initial therapy 41 days (IQR, 28.5–105.5) |

5-FC, flucytosine; AE, adverse event; dAmB, deoxycholate amphotericin B; L-AmB, liposomal amphotericin B; CM, cryptococcal meningitis; CSF, cerebrospinal fluid; EFA, early fungicidal activity; FLC, fluconazole; VOR, voriconazole

† Successful response defined according to reference [39]

**Table S3. Systematic review of publications evaluating combination antifungal therapy for invasive aspergilosis, 2003-2023**

| 1st author, year | Study design | Study period | Study population | N | Antifungal therapy | Treatment indication | Results - Primary endpoint | Safety outcomes | Pediatric patients | Comments |
|---|---|---|---|---|---|---|---|---|---|---|
| Elanjik al, 2003[40] | Case report | 2001 | 2F ALL | 1 | L-AmB 5 mg/kg/d + CAS 2 mg/kg x 1 then 1.5 mg/kg/d | Salvage therapy, proven IPA | Survived | Hypokemia with AmB | Y | |

| | | | | | | | | | | |
|---|---|---|---|---|---|---|---|---|---|---|
| Lujber, 2003[41] | Case report | 1999 | 25F immunocompetent | 1 | L-AmB 6-10 mg/kg/d + 5-FC 8 g/d | Salvage therapy, chronic invasive fungal rhinosinusitis with orbital and cerebral involvement | Survived | none | N | |
| Kontoyiannis, 2003[42] | Retrospective cohort study | 2001 | HM or HCT | 48 (n=23 proven or probable IA) | L-AmB 5 mg/kg/d + CAS | Primary or salvage therapy, proven, probable or possible IA | Success in 2/6 (33%) initial therapy cases and 3/17 (18%) salvage cases | Mild to moderate renal insufficiency in 15% | N | |
| Damaj, 2004[43] | Case report | NR | 57M AML | 1 | VOR + CAS | Salvage therapy, disseminated IA (lungs and CNS) | Survived | NR | N | IA initially progressed on dAmB monotherapy and dAmB + CAS combination |
| Marr, 2004[44] | Retrospective cohort study | 1997-2001 | HM or HCT | 47 | VOR (n=31) vs. VOR + CAS (n=16) | Salvage therapy proven or probable IA | Improved 3-month survival with VOR + CAS vs. VOR alone | | N | All pts received AmB or L-AmB for primary therapy |
| Rosen-Wolff, 2004[45] | Case report | NR | 13M CGD | 1 | LAmB 5 mg/kg/d + CAS 1 mg/kg/d | Primary therapy, proven IPA (*A. nidulans* and *A. fumigatus*) | Progressive infection requiring change in antifungal therapy to VOR monotherapy | Hypokemia with AmB | Y | Infection resolution on VOR monotherapy |
| Castagnola, 2004[46] | Case series | NR | 11M HCT | 1 | VOR + CAS 40 mg/m2/d loading, then 30 mg/m2/d | Salvage therapy, proven IA | Died | NR | Y | Death due to septic shock (Methicillin-resistant *Staphylococcus aureus*) |
| Elzi, 2005[47] | Case report | NR | 49M kidney transplant | 1 | ABLC 5 mg/kg/d + CAS 50 mg/d + 5-FC 3.5 g/d | Primary therapy, disseminated IA | Died | Leukopenia related to 5-FC | N | |
| Schuster, 2005[48, 49] | Case report | NR | 4M ALL | 1 | VOR + CAS 1 mg/kg/d | Salvage therapy, probable IPA | Survived | None | Y | |

| | | | | | | | | | | |
|---|---|---|---|---|---|---|---|---|---|---|
| Kontoyiannis, 2005 | Retrospective | 1993-2003 | HM or HCT | 112 | L-AmB 5 mg/kg/d (n=101), L-AmB + ITC (n=11) | Primary therapy proven or probable IA | No difference in response at end of primary therapy nor in 2-week or 4-week mortality between groups | | N | Excluded subjects receiving ITC capsules |
| Pancham, 2005[50] | Case series | NR | 14M HCT | 1 | L-AmB 5 mg/kg/d + CAS 50 mg/d | Salvage therapy, probable IPA (*A. fumigatus*) | Survived | None | Y | |
| Lehrnbecher, 2006[51] | Case report | NR | 10M HCT | 1 | L-AmB 5 mg/kg/d + VOR | Salvage therapy, proven intestinal IA | Survived | Elevated LFTs requiring temporary switch from VOR to CAS | Y | Laparotomy and partial small bowel resection performed |
| Denning, 2006[52] | Prospective, open-label, non-comparative, multicenter | 1998-2002 | HCT, HM, SOT, HIV/AIDS, Others | 225 | MFG 75 mg daily with optional dose escalation alone (n=34) or in combination with other agents (n=191, primarily L-AMB) | Primary therapy (n=29) or salvage therapy (n=196) proven or probable IA | Primary therapy: Favorable response (CR+PR) at end of therapy in 29.4% (combination) vs 50% (MFG alone) Salvage therapy: Favorable response in 34.5% (combination) vs 40.9% (MFG alone) | MFG well-tolerated alone or in combination | Y | |
| Maertens, 2006[53] | Prospective, open-label, non-comparative, multicenter | 2003-2004 | HCT, HM, Corticosteroids | 53 | CAS + ITC (n=7) or VOR (n=30) or dAmB 0.75-1.5 mg/kg/d (n=5) or L-AmB ≥ 5 mg/kg/d (n=11) | Salvage therapy proven or probable IA | Favorable response at end of therapy in 55% (50% CAS + polyene group and 56.8% CAS + triazole group) Survival at Day 84 - 55% | | N | Nine patients received combination therapy not including CAS as part of initial regimen |

| | | | | | | | | | | |
|---|---|---|---|---|---|---|---|---|---|---|
| Singh, 2006[54] | Prospective, observational, multicenter | 2003-2005 | SOT | 87 | VOR + CAS (n=40) vs historical control group lipid formulation AmB alone 5-7.4 mg/kg/d (n=47) | Primary therapy proven or probable IA | No difference in overall survival Day 90 in combination (67.5%) vs control groups (51%) | | N | Combination therapy associated with improved survival in subgroup with renal failure and in those with A. fumigatus infection; No correlation observed between in vitro antifungal interactions and clinical outcomes |
| Cesaro, 2007[55] | Prospective, multicenter surveillance study | 2002-2005 | Pediatric heme-onc or HCT | 40 | L-AmB 3-5 mg/kg/d plus CAS (n=18), VOR plus CAS (n=9), or both combinations sequentially (n=9) | Primary or salvage therapy proven or probable IA | Favorable response in 53%; 100-day survival probability 70% | Well-tolerated, no severe renal or liver impairment | Y | |
| Caillot, 2007[56] | Multicenter, randomized, open-label (Combistrat) | 2004-2005 | HM | 30 | L-AmB 3 mg/kg/d + CAS vs. L-AmB 10 mg/kg/d alone | Primary therapy proven or probable IA | Higher favorable response rate at end of therapy in combination (67%) vs. monotherapy group (27%) P = 0.028 | | Y | 23% of patients in high-dose monotherapy group experienced 2-fold increase in serum Cr |
| Meyer, 2007[57] | Case report | 2004 | 54M AML | 1 | VOR + dAmB 1.5 mg/kg/d | Salvage therapy, proven IPA | Progressive disease, requiring surgical resection | NR | N | Lower lobe resection required |
| Gubler, 2007[58] | Case report | NR | 43M cirrhosis | 1 | VOR 50-200 mg BID + CAS | Primary therapy, disseminated IA (lungs and CNS, *A. fumigatus*) | Survived | NR | N | |
| Panda, 2008[59] | Case series | NR | Adult immunocompetent | 6 | dAmB 1-1.5 mg/kg/d (gradual dose escalation) + ITC 300 mg x 2 doses, then 200 mg BID | Primary therapy, invasive fungal sinusitis with orbital +/- intra-cranial extension | 100% disease-free survival | NR | N | Surgical debridment in 1/6 patients |

| | | | | | | | | | | |
|---|---|---|---|---|---|---|---|---|---|---|
| Raad, 2008[60] | Single center compassionate use trial | 1994-2005 | HM or HCT | 143 | POS 800 mg/d suspension (n=53) vs. historical controls L-AmB ≥ 7.5 mg/kg/d alone (n=52) or with CAS (n=38) | Salvage therapy proven or probable IA | Higher favorable response rate in POS group (40%) vs L-AMB alone or with CAS (8% and 11%, respectively) | Higher rate of nephrotoxicity and hepatotoxicity in L-AmB alone and combination group | N | Most patients (~65%) received L-AmB 3-5 mg/kg/d for primary therapy; |
| Saijo, 2008[61] | Case report | NR | 56M alcoholic liver disease | 1 | MFG 300 mg/d + VOR 400 mg/d | Salvage therapy, probable IA (lung and CNS) | Survived | Liver dysfunction, reversed after VOR dose reduction | N | Initial therapy: MFG 150 mg/d |
| Matsuda, 2010[62] | Case report | 2005 | 62M interstitial pneumonia on systemic immunosuppression | 1 | VOR 300 mg BID + MFG 150 mg/d | Salvage therapy, proven *Aspergillus* empyema | Clinical, radiographic, and mycological resolution | Elevated LFTs requiring VOR dose reduction | N | |
| Yamada, 2010[63] | Case report | 2007 | 45M AML | 1 | L-AmB 200 m/d + MFG 300 mg/d, followed by VOR 400 mg/d + MFG | Salvage therapy, *Aspergillus* liver abscess | Clinical and radiographic resolution | Elevated serum creatinine attributed to L-AmB | N | |
| Aoki, 2011[64] | Case report | 2007-2008 | 49F AML, 48F AML | 1 | Case 1: VOR + MFG 150 mg/d; Case 2: VOR 400 mg/d + MFG 150 mg/d | Salvage therapy, probable IPA | Survived | None | N | |
| Beiras-Fernandez, 2011[65] | Case report | NR | 45M heart transplant | 1 | VOR 400 mg/d + MFG 100 mg/d | Primary therapy, IPA | Clinical and radiographic resolution | None | N | |
| Kerl, 2011[66] | Case report | NR | 5M HCT | 1 | VOR 10 mg/kg BID + CAS 50 mg/d | Primary therapy, cutaneous IA | Resolved | Elevated liver transaminases and facial rash possibly related to VOR | Y | Surgical debridement performed |

| | | | | | | | | | | |
|---|---|---|---|---|---|---|---|---|---|---|
| Lazaro, 2011[67] | Case report | 2009 | 40M lung transplant | 1 | VOR 400 mg BID x 2 doses, then 300 mg BID + AFG | Primary therapy, Aspergillus fumigatus infectious endocarditis | Died | NR | N | |
| Dupouey, 2012[68] | Case series | NR | Aortic valve surgery | 3 | Case 1: VOR + CAS, Case 2: dAmB 70 mg/d + 5-FC 10 g/d, Case 3: CAS + L-AmB 5 mg/kg/d + VOR | Primary therapy, *A. flavus* infectious endocarditis | Case 1: survived, Cases 2 and 3: died | NR | N | |
| Doring, 2012[69] | Case report | 2006 | 18M ALL and HCT x 3 | 1 | L-AmB 3 mg/kg/d + CAS | Primary therapy, probable IPA | Clinical and radiographic improvement, then worsening after switch to VOR | Elevated blood urea nitrogen possibly related to L-AmB | N | |
| Rojas, 2012[70] | Retrospective, multicenter, observational study | 2005-2010 | HM or HCT | 61 (n=49 with IA) | L-AmB 3 mg/kg/d + CAS (n=18), L-AmB 3 mg/kg/d + VOR or POS 400 mg q 12 h (n=14), or VOR + EC (n=17) | Initial therapy, proven or probable IA | Favorable response and 12-week survival, respectively, in 13/18 (72%) L-AmB + CAS group, 8/14 (57%)L-AmB + VOR or POS group, 11/17 (65%) VOR + EC group | Mild elevated LFTs (VOR) and mild increase serum creatinine (L-AmB) | Y | Pediatric VOR dosing 7 mg/kg q12 h |
| Pillai, 2013[71] | Case report | NR | 47M pacemaker surgery | 1 | L-AmB 5 mg/kg/d + VOR 200 BID | Primary therapy, pacemaker lead *Aspergillus* endocarditis | Survived | None | N | |
| Demir, 2015[72] | Case report | NR | 36M HCT | 1 | L-AmB 3 mg/kg/d + CAS | Salvage therapy, disseminated IA (lungs, heart, skin, CNS) | Died | NR | N | |

| | | | | | | | | | | |
|---|---|---|---|---|---|---|---|---|---|---|
| Marr, 2015[73] | Randomized, double-blind, placebo-controlled, multicenter | 2008-2011 | HM or HCT | 277 | VOR alone (n=142) or with AFG (n=135) | Primary therapy proven or probable IA | No difference in 6-week all-cause mortality between groups: 19.3% (VOR + AFG) vs. 27.5% (VOR) p=0.087 | Similar rates of TEAEs except more hepatobiliary disorders in combination group (12.7%) than monotherapy group (8.4%) | N | Post-hoc subroup analysis of probable IA cases (n=218) found lower 6-week mortality in combination (15.7%) vs monotherapy (27.3%) groups; p=0.037 Therapeutic drug monitoring not performed |
| Raad, 2015[74] | Retrospective, single-center | 1998-2010 | HM | 138 | CAS alone (n=32), VOR alone (n=62) or with CAS (n=68) | Primary or salvage therapy proven or probable IA | No difference in favorable response rates among the 3 groups for primary or salvage therapy No difference in IA-associated mortality between VOR monotherapy and VOR + CAS groups for primary or salvage therapy | Primary therapy: no difference in adverse events between groups Salvage therapy: More adverse events in combination group (37%) vs CAS alone (6%) | Y | |
| Rofaiel, 2016[75] | Case report | NR | 64F acute leukemia | 1 | L-AmB 5 mg/kg/d + 5-FC 25 mg/kg q 6 h | Primary therapy, *A. fumigatus* native valve endocarditis | Died | NR | N | |
| Nicolé, 2017[76] | Case report | 2015 | 70M cryoglobulinemic vasculitis on corticosteroids | 1 | VOR 200 mg q 12 h + CAS | Salvage therapy, *Aspergillus* thyroiditis | Persistent thyroid abscesses requiring total thyroidectomy | Liver toxicity on VOR | N | |
| Chen, 2017[77] | Case report | NR | 16M solid tumor | 1 | VOR 11 mg/kg q 12 h + CAS 100 mg/d | Salvage therapy, CNS aspergillosis | Resolved | Elevated liver transaminases | Y | |

| | | | | | | | | | | |
|---|---|---|---|---|---|---|---|---|---|---|
| Lee, 2019[78] | Subgroup analysis from RCT (ref Marr) | 2008-2011 | HM or HCT (Korean patients only) | 40 | VOR alone (n=17) or with AFG (n=23) | Primary therapy proven or probable IA | Week 6 all-cause mortality numerically lower in combination arm (17.6%) than monotherapy arm (41.2%), a more marked reduction in mortality as compared to non-Koreans | No difference in TEAEs between groups | N | |
| Calvo-Cano, 2020[79] | Case report | | 56M kidney transplant | 1 | L-AmB 8 mg/kg/d + 5-FC 3 g 3x/d + inhaled AmB | Salvage therapy, azole-resistant disseminated IA (*A. parafelis*, lungs and CNS) | Progressive disease requiring neurosurgery (lobectomy) | | N | Following neurosurgery, switched to MFG 350 3x/week + POS DR 300 mg/d maintenance therapy |
| Furudate, 2020[80] | Case report | NR | Preterm infant | 1 | VOR 4.5-9 mg/kg q 12 h + L-AmB 5 mg/kg/d | Salvage therapy, IPA and *Aspergillus* ventriculitis | Resolved | Elevated LFTs requiring temporary discontinuation of VOR | Y | Endoscopic intraventricular lavage also performed |
| Lennard, 2020[81] | Case report | 2016 | 62M aortic valve replacement | 1 | L-AmB 5 mg/kg/d + VOR | Primary therapy, *Aspergillus fumigatus* prosthetic valve endocarditis | Resolved | NR | N | Aortic root debridement and aortic valve replacement performed |
| Mendoza, 2020[82] | Case report | NR | 56F HCT | 1 | L-AmB 5 mg/kg/d + MFG 150 mg/d + TRB 500 mg q 12 h | Salvage therapy, disseminated IA (*A. calidoustus*) | Clinical and mycological response (downtrending serum GM) | AKI requiring switch from L-AmB to ISA | N | In vitro synergy testing performed but methodology not specified: synergy between ISA and TRB (MICs: 0.5 + 0.03 µg/mL, respectively) but no synergy (indifferent) for MFG + ISA combination (MICs: ≤ 0.06 + 2 µg/mL, respectively) |

| | | | | | | | | | | |
|---|---|---|---|---|---|---|---|---|---|---|
| Camargo, 2022[83] | Case report | NR | 40M HCT and GVHD | 1 | L-AmB 5 mg/kg/d + MFG 100-150 mg/d + ISA + TRB 500 mg BID + Fosmanogepix 1 g q 12 h x 2 doses then 600 mg/d | Primary therapy, disseminated IA (*A. calidoustus*) | Resolved | NR | N | Continued on fosmanogepix 400 mg BID alone as of last follow-up 324 days after IA diagnosis. In vitro synergy testing performed but methodology not specified: synergy between AmB + MFG (≤ 0.03), AmB + POS (≤ 0.03), AmB + VOR (≤ 0.03), AmB + TRB (≤ 0.015), AmB + ISA (≤ 0.03), ISA + TRB (≤ 0.03). |
| Nakaya, 2023[84] | Case report | NR | 29F HCT and GVHD | 1 | VOR 300-600 mg/d + CAS 70 mg/d --> progressive disease --> ITC 200 mg/d + MFG 150 mg/d -> progressive disease --> POS 200-600 mg/d + L-AmB 5 mg/kg/d | Salvage therapy, COVID-19 associated pulmonary aspergillosis (*A. fumigatus*) | Resolved | Hypokemia with AmB | N | |

5-FC, flucytosine; ABLC, lipid complex amphotericin B; AKI, acute kidney injury; ALL, acute lymphoblastic leukemia; CAS, caspofungin; CGD, chronic granulomatous disease; CNS, central nervous system; dAmB, deoxycholate amphotericin B; GM, galactomannan; GVHD, graft-versus-host disease; HCT, hematopoietic cell transplantation; HM, hematologic malignancy; ITC, itraconazole; IPA, invasive pulmonary aspergillosis; L-AmB, liposomal amphotericin B; LFTs, liver function tests; MFG, micafungin; NR, not reported; PVE, prosthetic valve endocarditis; SAE, serious adverse event; TRB, terbinafine

† Antifungal dosages unless otherwise specified: AFG 200 mg x 1 dose followed by 100 mg/d, CAS 70 mg x 1 dose followed by 50 mg/d, ISA 372 mg q 8 h x 6 doses followed by 372 mg/d, ITC 200 mg BID x 2 days followed by 200 mg/d, VOR 6 mg/kg q 12 h x 2 doses followed by 4 mg/kg q 12 h or 200 mg q 12 h

**Table S4. Systematic review of publications evaluating combination antifungal therapy for mucormycosis, 2003-2023**

| 1st author, year | Study design | Study period | Study population | N | Antifungal therapy† | Treatment indication | Results - Primary endpoint | Safety outcomes | Pediatric patients | Comments |
|---|---|---|---|---|---|---|---|---|---|---|
| Rickerts, 2006[85] | Case report | 2005 | 46F AML | 1 | L-AmB 5 mg/kg/d + POS 400 mg BID | Salvage, disseminated IM *(Rhizomucor* spp.) | Survived | NR | N | |
| Reed, 2008[86] | Retrospective cohort study, two centers | 1994-2006 | ROCM (83% diabetic, 34% cancer, 12% neutropenia, 10% transplant) | 41 | dAmB 1 mg/kg/d (n=15), ABLC 5 mg/kg/d (n=17), L-AmB 5 mg/kg/d (n=2); CAS + ABLC (n=5), CAS + L-AmB (n=2) | Primary therapy proven IM | Alive and not in hospital care 30 days after hospital discharge: 100% combination therapy vs. 45% monotherapy, P = 0.02 | Doubling of serum Cr in 56% during therapy with no difference between polyene agent | Y | Among patients with CNS disease, primary endpoint met in 4/4 (100%) combination group vs. 25% monotherapy group |
| Lebeau, 2010[87] | Case report | NR | 46F AML | 1 | L-AmB 5 mg/kg/d + POS 400 mg BID | Primary, disseminated IM (*R. microsporus*) | Survived | Mild chronic renal insufficiency | N | |
| Roux, 2010[88] | Case report | NR | 13F metastatic osteosarcoma | 1 | L-AmB 10 mg/kg/d + POS; CAS added 1 month later | Primary, disseminated IM (*Absidia corymbifera*) | Survived | NR | Y | POS dose not specified |
| Ogawa, 2012[89] | Case report | NR | 70M diabetes | 1 | L-AmB 10 mg/kg/d + MFG 6 mg/kg/d | Primary, invasive fungal sinusitis ( R. oryzae) | Survived | NR | N | |
| Bourke, 2012[90] | Case report | 2007 | 53M heart transplant | 1 | L-AmB 7.5 mg/kg/d + POS 200 mg QID | Salvage, invasive fungal sinusitis *(Rhizopus* spp.) | Survived | Acute kidney injury | N | |

| | | | | | | | | | | |
|---|---|---|---|---|---|---|---|---|---|---|
| Guymer, 2013[91] | Case report | 2011 | 11F T-ALL | 1 | L-AmB 10 mg/kg/d + CAS 70 mg/m2 loading dose, then 50 mg/m2/d, then L-AmB 5 mg/kg/d + POS beginning day 82 | Primary, disseminated IM (*Lichtheimia corymbifera*) | Survived | NR | Y | Aggressive surgical debridement performed; POS dose not specified |
| Pagano, 2013[92] | Analysis of SEIFEM and Fungiscope registries | 2007-2012 | HM | 32 | ABLC + POS (n=5), L-AmB 3 mg/kg + POS (n=10), L-AmB ≥ 5 mg/kg/d + POS (n=17) | Primary or salvage therapy proven or probable IM | Clinical improvement (CR or PR) in 56% and IM-related death in 28% at median 3 month follow-up | No patients stopped AF treatment due to drug toxicity | NR | Salvage indication for 29 (91%) patients, nearly all started on lipid formulation AmB then POS added; POS dose 800 mg/kg/d in 28 (88%) patients |
| Klimko, 2014[93] | Case report | 2010 | 53F AML | 1 | dAmB 1.5 mg/kg/d + CAS | Salvage, musculoskeletal (*Lichtheimia corymbifera*) | Survived | NR | N | |
| Di Carlo, 2014[94] | Case report | 2013 | 70F diabetes | 1 | L-AmB 3 mg/kg/d + POS 400 mg BID | Salvage, ROCM | Survived | NR | N | |
| Kyvernitakis, 2016[95] | Retrospective cohort study, single-center | 1994-2014 | HM and HCT | 106 | Monotherapy, n=47: L-AmB < 5 to ≥ 7.5 mg/kg/d (87%) or POS (13%); Combination therapy, n=59: L-AmB + POS (27%), L-AmB + EC (46%), or L-AmB + POS + EC (27%) | Primary therapy of IM | No difference in propensity score-adjusted probability of 6-week mortality between treatment groups (56% monotherapy vs. 60% combination, P = 0.71) | NR | N | Initial combination therapy also had no impact on 6-week survival based on mortality risk score, nor on overall survival at 12 weeks; POS dose not specified |
| Ville, 2016[96] | Case report | NR | 60M kidney transplant | 1 | L-AmB 10 mg/kg/d + POS 400 mg BID | Primary, disseminated IM (*R. microsporus*) | Died | NR | N | |

| | | | | | | | | | | |
|---|---|---|---|---|---|---|---|---|---|---|
| Al Hassan, 2020[97] | Case series | NR | Diabetes | 2 | L-AmB 10 mg/kg/d + POS 200 mg QID | Primary, ROCM | Died | NR | N | |
| Risum, 2020[98] | Retrospective, single-center case series | 2012-2019 | HM | 13 | Initial monotherapy, n=4: L-AmB 3-5 mg/kg/d (n=3) or POS (n=1); initial combination therapy, n=9: L-AmB + POS (n=5), L-AmB + POS + TRB (n=1), L-AmB + ISA (n=2), L-AmB + ISA + TRB (n=1) | Primary therapy of proven IM | 3-month survival: 7/9 combination patients and 4/4 monotherapy patients; cure achieved in 10 patients (6/9 combination and 4/4 montherapy) | NR | Y | Nine patients received concomitant topical AF therapy; POS dose 300 mg/d in 5/7 patients; TRB dose 4 mg/kg (n=1) and 1-2 g (n=1) |
| Salehi, 2020[99] | Case report | NR | 35F lymphoma | 1 | L-AmB 300 mg + POS 200mg QID | Primary, ROCM | Survived | NR | N | |
| Miller, 2021[100] | Retrospective cohort study, multicenter | 2007-2017 | HM and HCT | 64 | Initial monotherapy, n=33: Lipid formulation AmB (Lip-AmB) 5-10 mg/kg/d alone (n=28), POS alone (n=3), ISA alone (n=2); initial combination therapy, n=31: Lip-AmB + POS (n=16) or EC (n=8) or ISA (n=5) or POS + EC (n=2) | Primary therapy proven or probable IM | No difference in treatment failure or 30-day mortality with Lip-AmB alone (64% and 43%, respectively) vs Lip-AmB + POS or ISA (43% and 28%, respectively), P > 0.1 | NR | N | |

| | | | | | | | | | | |
|---|---|---|---|---|---|---|---|---|---|---|
| Eubank, 2021[101] | Case report | NR | Liver transplant | 3 | Case 1: ISA + L-AmB 3 mg/kg/d, then L-AmB + POS 300 mg/d + MFG 150 mg/d; Case 2: L-AmB 3 mg/kg/d + ISA + MFG 150 mg/d, then added terbinafine 500 mg BID and L-AmB irrigation; Case 3: L-AmB 7.5 mg/kg/d + POS 300 mg BID + MFG 150 mg/d | Primary therapy *(Rhizopus* spp.) | Survived | NR | N | Infection sites: musculoskeletal, intra-abdominal, sinus |
| Beaver, 2021[102] | Case report | NR | 47M immunocompetent | 1 | L-AmB 10 mg/kg/d + ISA x 25 days, then added MFG 100 mg + topical AmB | Primary, sino-orbital IM *(Rhizomucor* spp.) | Survived | Acute kidney injury | N | |
| Khera, 2021[103] | Case report | NR | 6M B-ALL | 1 | L-AmB 8 mg/kg/d + CAS 50 mg/m2 | Salvage, pulmonary | Survived | NR | Y | |
| Kaushal, 2022[104] | Case series | 2017-2019 | Diabetes | 2 | Case 1: L-AmB 10.15 g/d + POS 126.40 g; Case 2: L-AmB 4 g/d + POS 2 g | Salvage, invasive fungal sinusitis, ROCM | Case 1: survived; Case 2: died | none | N | |
| Loeffen, 2022[105] | Retrospective, single-center case series | 2018-2021 | HM, pediatric | 10 (n=8 proven or probable cases) | Initial therapy: L-AmB 10 mg/kd/d + ISA (n=4), ABLC 7.5-10 mg/kg/d + ISA (n=2), L-AmB 5 mg/kg/d + POS (n=1) | Primary therapy proven, probable, possible IM | Two IM-related deaths, 1 death due to underlying HM, 5 patients alive with 4-22 months of follow-up | 5 patients with reversible nephrotoxicity on L-AmB | Y | |

ABLC, lipid complex amphotericin B; ALL, acute lymphoblastic leukemia; AML, acute myelogenous leukemia; dAmB, deoxycholate amphotericin B; HM, hematologic malignancy; ISA, isavuconazole; L-AmB: liposomal amphotericin B; NR: not reported; POS, posaconazole; ROCM, rhinorbital cerebral mucormycosis; SEIFEM: Sorveglianza Epidemiologica Infezioni Fungine Emopatie Maligne; T-ALL, T-cell acute lymphoblastic leukemia; TRB, terbinafine

† Antifungal dosages unless otherwise specified: CAS 70 mg x 1 dose followed by 50 mg/d, ISA 372 mg q 8 h x 6 doses followed by 372 mg/d
