## Supplemental Figure 1 for "CAF to the rescue! Potential and challenges of combination antifungal therapy for reducing morbidity and mortality in hospitalized patients with serious fungal infections"

**Supplementary Figure 1.** Literature analysis of publications pertaining to laboratory testing of antifungal drugs in combinations against yeasts and molds

**Laboratory Methods**

<u>Search Criteria: PubMed (1984-2024)</u>

| | |
|---|---|
| Generic Search Terms (All fields) | |
| Activity (Activities) | 6,327,959 |
| Testing | 3,371,750 |
| Combination | 2,685,343 |
| In vitro | 1,927,465 |
| Susceptibility | 642,043 |
| Antifungal | 232,801 |
| Antifungal combination | 32,729 |
| Broth dilution | 5,002 |
| Antifungal combination testing | 6,168 |
| In vitro antifungal combination | 5,269 |
| In vitro activity antifungal combination | 3,293 |
| Antifungal combination susceptibility | 1,873 |
| Antifungal combination susceptibility testing | 1,117 |
| Antifungal combination testing broth dilution | 76 |
| Antifungal combination susceptibility testing broth dilution | 37 |
| In vitro activity antifungal combination broth dilution | 29 |

<u>Google scholar</u>

| | |
|---|---|
| Antifungal combination testing (exact phrase) | 50 (10) |

<u>76+37+29 publications in English examined for the inclusion of approved antifungal drugs and QC strains = 22 studies</u>
