## Appendix for "CAF to the rescue! Potential and challenges of combination antifungal therapy for reducing morbidity and mortality in hospitalized patients with serious fungal infections"

Keywords used for the PubMed Search strategy:

#1

(("candida"[Title/Abstract] OR "candidiasis"[Title/Abstract]) AND "combination therapy"[Title/Abstract] AND 2003/01/01:3000/12/31[Date - Publication]) OR (("candida"[Title/Abstract] OR "candidiasis"[Title/Abstract]) AND "combination antifungal therapy"[Title/Abstract] AND 2003/01/01:3000/12/31[Date - Publication])

#2

(("cryptococcus"[Title/Abstract] OR "cryptococcal"[Title/Abstract] OR "cryptococcosis"[Title/Abstract]) AND "combination therapy"[Title/Abstract] AND 2003/01/01:3000/12/31[Date - Publication]) OR (("cryptococcus"[Title/Abstract] OR "cryptococcal"[Title/Abstract] OR "cryptococcosis"[Title/Abstract]) AND "combination antifungal therapy"[Title/Abstract] AND 2003/01/01:3000/12/31[Date - Publication])

#3

(("aspergillus"[Title/Abstract] OR "aspergillosis"[Title/Abstract]) AND "combination therapy"[Title/Abstract] AND 2003/01/01:3000/12/31[Date - Publication]) OR (("aspergillus"[Title/Abstract] OR "aspergillosis"[Title/Abstract]) AND "combination antifungal therapy"[Title/Abstract] AND 2003/01/01:3000/12/31[Date - Publication])

#4

(("mucormycosis"[Title/Abstract] OR "mucorales"[Title/Abstract] OR "zygomycetes"[Title/Abstract] OR "zygomycosis"[Title/Abstract]) AND "combination therapy"[Title/Abstract] AND 2003/01/01:3000/12/31[Date - Publication]) OR (("mucormycosis"[Title/Abstract] OR "mucorales"[Title/Abstract] OR "zygomycetes"[Title/Abstract] OR "zygomycosis"[Title/Abstract]) AND "combination antifungal therapy"[Title/Abstract] AND 2003/01/01:3000/12/31[Date - Publication])

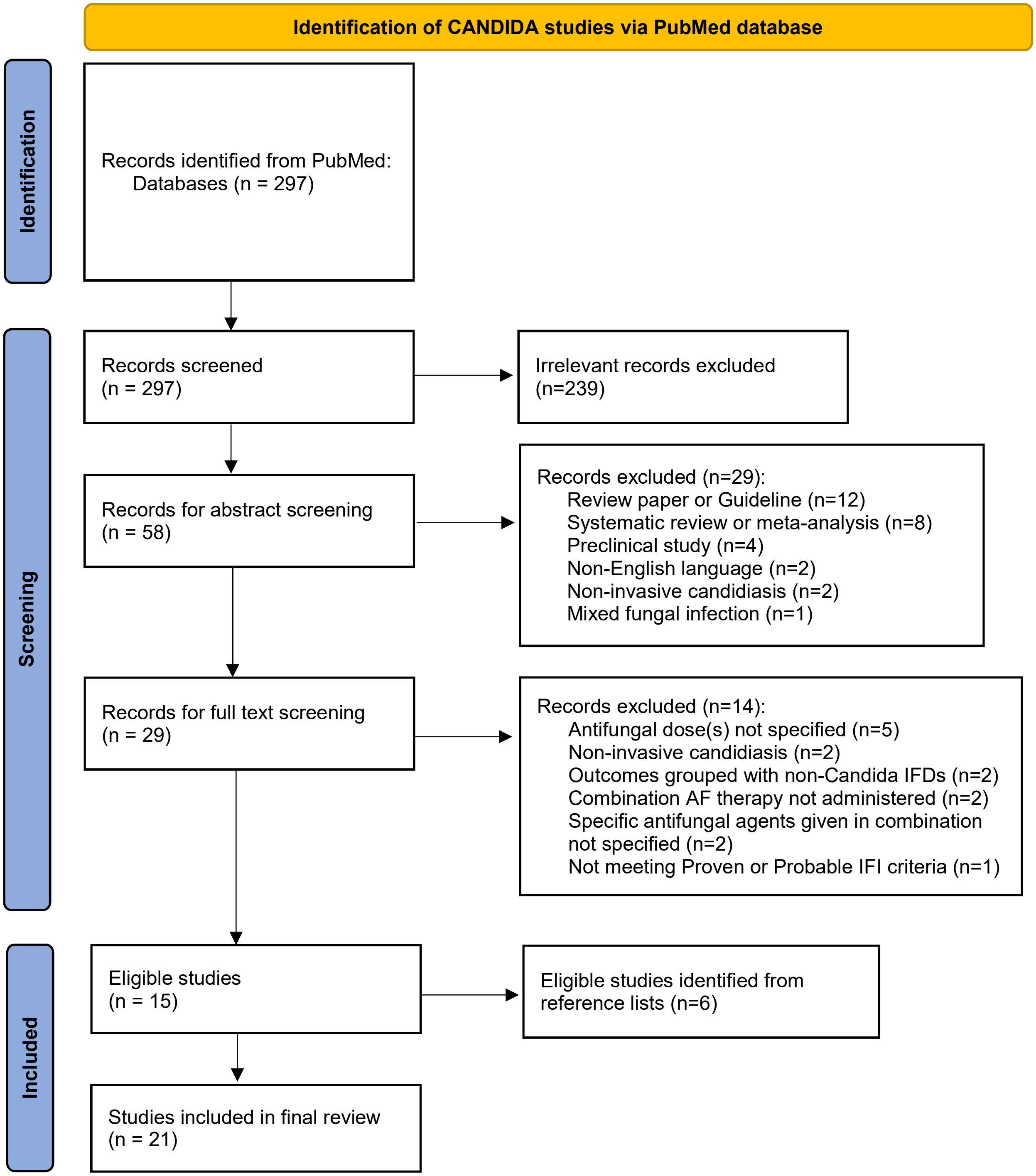
Identification of CANDIDA studies via PubMed database
Identification
Records identified from PubMed: Databases (n = 297)
Screening
Records screened (n = 297)
Irrelevant records excluded (n=239)
Records for abstract screening (n = 58)
Records excluded (n=29):
Review paper or Guideline (n=12)
Systematic review or meta-analysis (n=8)
Preclinical study (n=4)
Non-English language (n=2)
Non-invasive candidiasis (n=2)
Mixed fungal infection (n=1)
Records for full text screening (n = 29)
Records excluded (n=14):
Antifungal dose(s) not specified (n=5)
Non-invasive candidiasis (n=2)
Outcomes grouped with non-Candida IFDs (n=2)
Combination AF therapy not administered (n=2)
Specific antifungal agents given in combination not specified (n=2)
Not meeting Proven or Probable IFI criteria (n=1)
Included
Eligible studies (n = 15)
Eligible studies identified from reference lists (n=6)
Studies included in final review (n = 21)

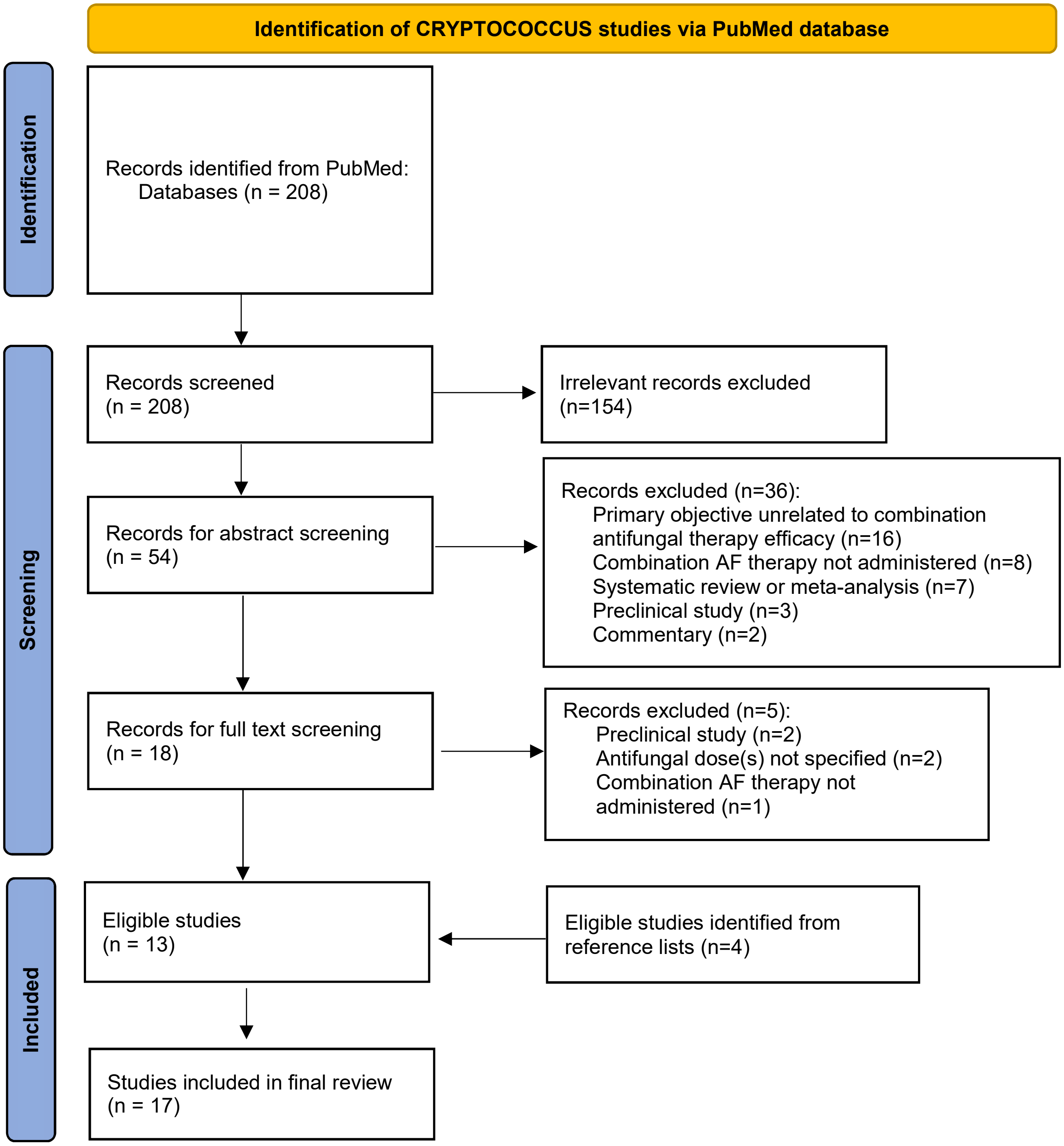
Identification of CRYPTOCOCCUS studies via PubMed database
Identification
Records identified from PubMed:
Databases (n = 208)
Screening
Records screened
(n = 208)
Irrelevant records excluded
(n=154)
Records for abstract screening
(n = 54)
Records excluded (n=36):
Primary objective unrelated to combination antifungal therapy efficacy (n=16)
Combination AF therapy not administered (n=8)
Systematic review or meta-analysis (n=7)
Preclinical study (n=3)
Commentary (n=2)
Records for full text screening
(n = 18)
Records excluded (n=5):
Preclinical study (n=2)
Antifungal dose(s) not specified (n=2)
Combination AF therapy not administered (n=1)
Included
Eligible studies
(n = 13)
Eligible studies identified from reference lists (n=4)
Studies included in final review
(n = 17)

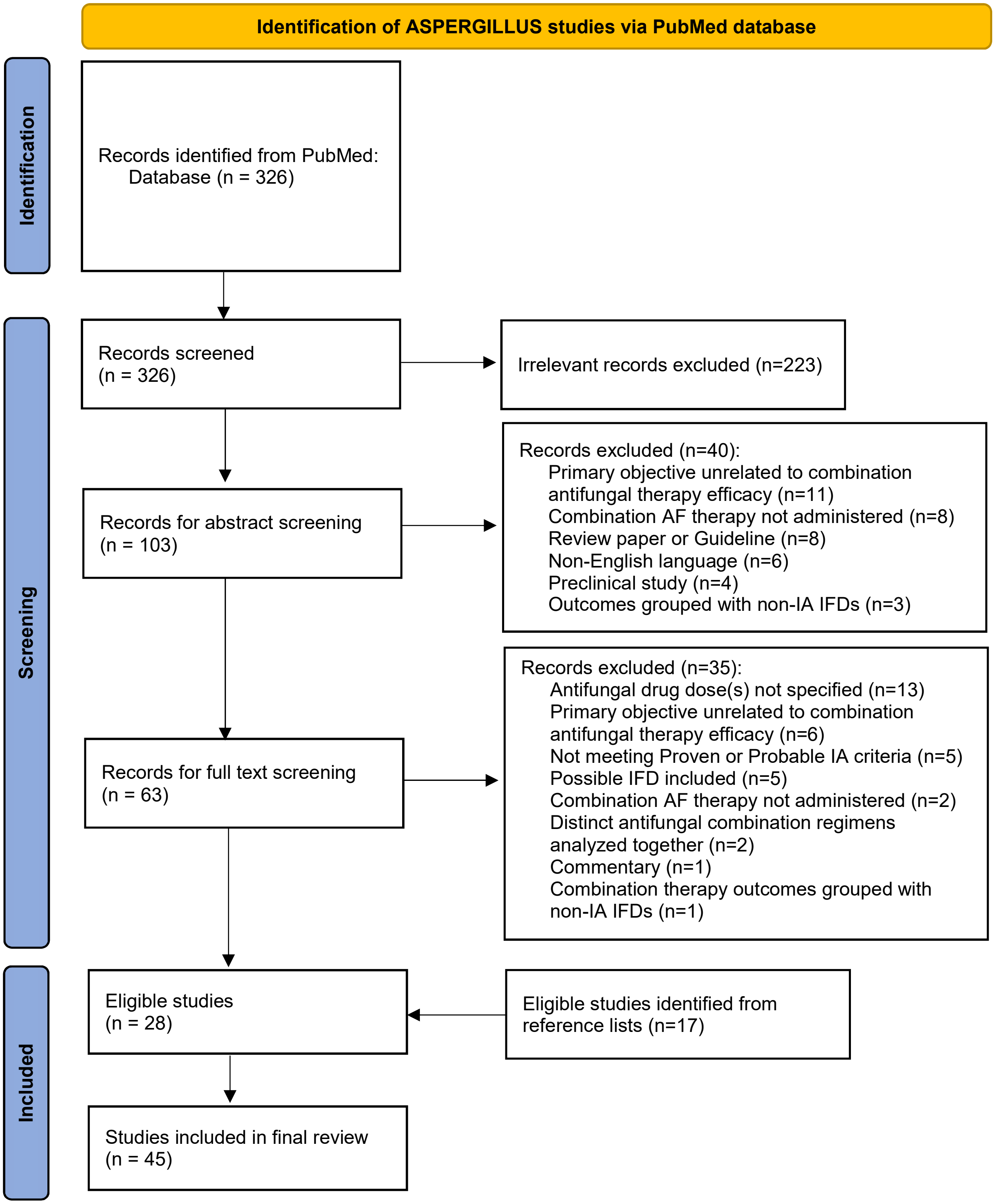
Identification of ASPERGILLUS studies via PubMed database
Identification
Screening
Included
Records identified from PubMed: Database (n = 326)
Records screened (n = 326)
Irrelevant records excluded (n=223)
Records for abstract screening (n = 103)
Records excluded (n=40):
Primary objective unrelated to combination antifungal therapy efficacy (n=11)
Combination AF therapy not administered (n=8)
Review paper or Guideline (n=8)
Non-English language (n=6)
Preclinical study (n=4)
Outcomes grouped with non-IA IFDs (n=3)
Records for full text screening (n = 63)
Records excluded (n=35):
Antifungal drug dose(s) not specified (n=13)
Primary objective unrelated to combination antifungal therapy efficacy (n=6)
Not meeting Proven or Probable IA criteria (n=5)
Possible IFD included (n=5)
Combination AF therapy not administered (n=2)
Distinct antifungal combination regimens analyzed together (n=2)
Commentary (n=1)
Combination therapy outcomes grouped with non-IA IFDs (n=1)
Eligible studies (n = 28)
Eligible studies identified from reference lists (n=17)
Studies included in final review (n = 45)

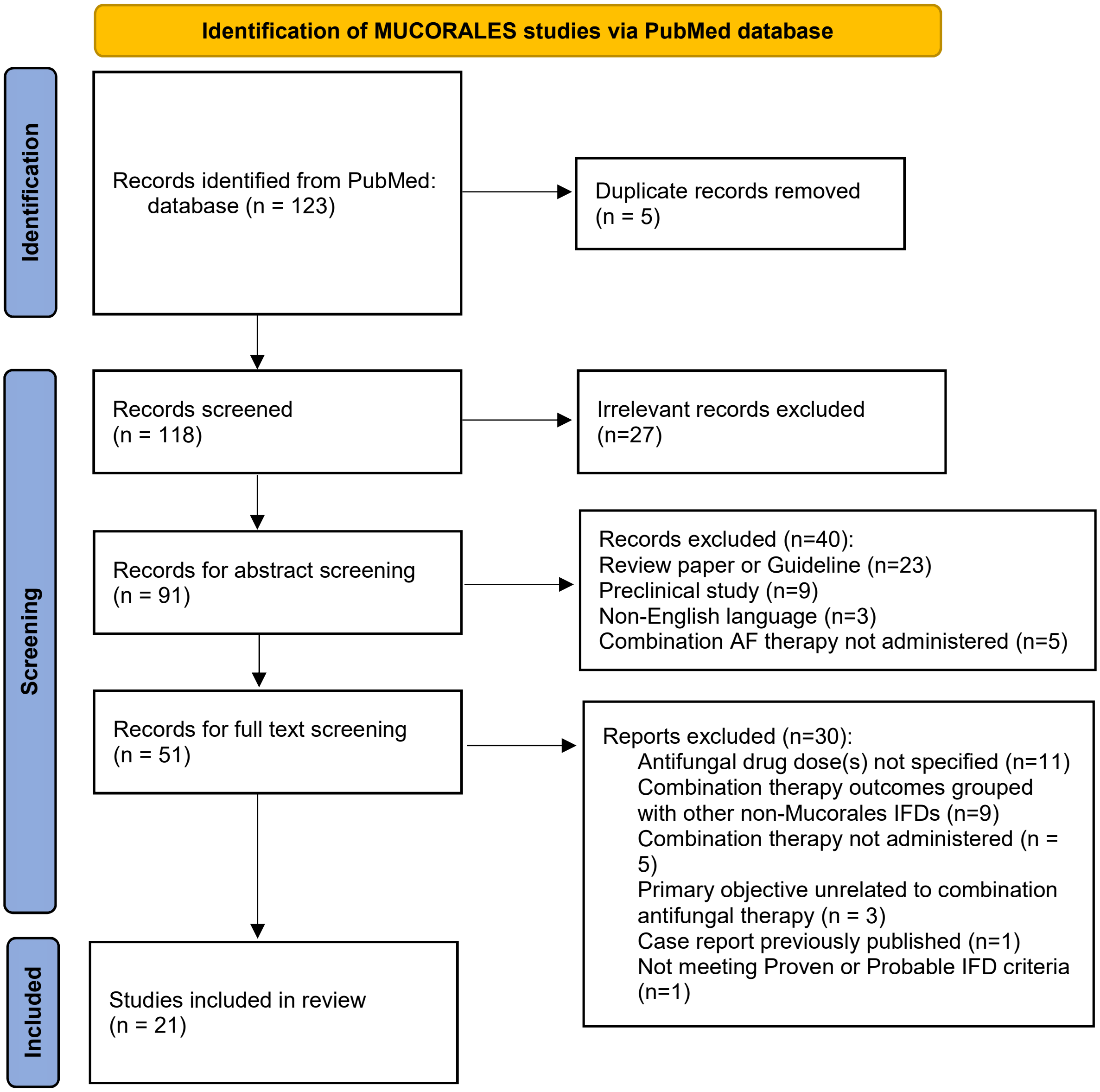
Identification of MUCORALES studies via PubMed database
Identification
Screening
Included
Records identified from PubMed: database (n = 123)
Duplicate records removed (n = 5)
Records screened (n = 118)
Irrelevant records excluded (n=27)
Records for abstract screening (n = 91)
Records excluded (n=40):
Review paper or Guideline (n=23)
Preclinical study (n=9)
Non-English language (n=3)
Combination AF therapy not administered (n=5)
Records for full text screening (n = 51)
Reports excluded (n=30):
Antifungal drug dose(s) not specified (n=11)
Combination therapy outcomes grouped with other non-Mucorales IFDs (n=9)
Combination therapy not administered (n = 5)
Primary objective unrelated to combination antifungal therapy (n = 3)
Case report previously published (n=1)
Not meeting Proven or Probable IFD criteria (n=1)
Studies included in review (n = 21)
